# A systematic review and meta-analysis of cancer patients affected by a novel coronavirus

**DOI:** 10.1101/2020.05.27.20115303

**Authors:** BP Venkatesulu, VT Chandrasekar, P. Girdhar, P. Advani, A. Sharma, T. Elumalai, C. Hsieh, HI. Elghazawy, V. Verma, S. Krishnan

**Affiliations:** Transitional year residency, Department of Internal Medicine, Henry Ford Hospital, Detroit, MI 48202, United States.; Department of Gastroenterology and Hepatology, University of Kansas Medical Center, Kansas City, KS 66160, United States; Department of Radiation Oncology, All India Institute of Medical Sciences, New Delhi, India; Radiation Epidemiology Branch, Division of Cancer Epidemiology and Genetics, National Institutes of Health, National Cancer Institute, Bethesda, MD 20892, United States; Department of Experimental Radiation Oncology, The University of Texas MD Anderson Cancer Center, Houston, TX 77030, United States; Department of Clinical Oncology, The Christie NHS Foundation Trust, Manchester United Kingdom; Department of Radiation Oncology, Institute for Radiological Research, Chang Gung Memorial Hospital at Linkou and Chang Gung University, Taoyuan City, Taiwan; Graduate School of Biomedical Sciences, The University of Texas Health Science Center at Houston and The University of Texas MD Anderson Cancer Center, Houston, TX 77030, United States; Department of Clinical Oncology, Faculty of Medicine, Ain Shams University, Abbaseya, Cairo, Egypt; Department of Radiation Oncology, Allegheny General Hospital, Pittsburgh, Pennsylvania; Department of Radiation Oncology, Mayo Clinic Florida, 4500 San Pablo Road S, Mayo 1N, Jacksonville, FL 32224, USA

**Keywords:** COVID-19, meta-analysis, cancer;, chemotherapy, radiotherapy

## Abstract

**Background:** Cancer patients with COVID-19 disease have been reported to have double the case fatality rate of the general population.

**Materials and methods:** A systematic search of PubMed/MEDLINE, Embase, Cochrane Central, Google Scholar, and MedRxiv was done for studies on cancer patients with COVID-19. Pooled proportions were calculated for categorical variables.Odds ratio and forest plots were constructed for both primary and secondary outcomes. The random-effects model was used to account for heterogeneity between studies.

**Results:** This systematic review of 31 studies and meta-analysis of 181,323 patients from 26 studies involving 23,736 cancer patients is the largest meta-analysis to the best of our knowledge assessing outcomes in cancer patients affected by COVID-19. Our meta-analysis shows that cancer patients with COVID-19 have a higher likelihood of death (odds ratio, OR 2.54), which was largely driven by mortality among patients in China. Cancer patients were more likely to be intubated, although ICU admission rates were not statistically significant. Among cancer subtypes, the mortality was highest in hematological malignancies (OR 2.43) followed by lung cancer (OR 1.8). There was no association between receipt of a particular type of oncologic therapy and mortality. Our study showed that cancer patients affected by COVID-19 are a decade older than the normal population and have a higher proportion of co-morbidities. There was insufficient data to assess the association of COVID-directed therapy and survival outcomes in cancer patients. Despite the heterogeneity of studies and inconsistencies in reported variables and outcomes, these data could guide clinical practice and oncologic care during this unprecedented global health pandemic.

**Conclusion:** Cancer patients with COVID-19 disease are at increased risk of mortality and morbidity. A more nuanced understanding of the interaction between cancer-directed therapies and COVID-19-directed therapies is needed. This will require uniform prospective recording of data, possibly in multi-institutional registry databases.

## Introduction

Severe acute respiratory syndrome-related coronavirus 2 (SARS-CoV-2) is a novel beta-coronavirus, and is the causative agent of coronavirus disease 2019 (COVID-19)[1]. COVID-19 has caused an unprecedented global health pandemic, with more than 5 million cases and 0.33 million deaths reported worldwide (at the time of writing)[2]. Worldwide data suggest that there are around 18 million new cancer patients every year, with around 43 million patients living with a cancer diagnosis within the past 5 years [3,4]. A systematic review showed a pooled prevalence of cancer patients with COVID-19 to be around 2.0%[5]. However, cancer patients have been reported to have double the case fatality rates as compared to the general population[6]. The majority of cancer patients tend to be older, have multiple preexisting comorbidities, and are immunosuppressed from numerous causes[7]. Moreover, owing to oncologic interventions and follow-up thereof, time spent in the hospital as well as interaction with healthcare providers may further increase the proclivity to develop infections. For instance, radiotherapy requires multiple visits to the hospital due to its fractionated nature of the treatment and has been known to deplete circulating and resident T lymphocyte populations[8]. Since the main pathophysiologic driver of mortality in COVID-19 is the cytokine storm and macrophage activation, immunotherapy agents might augment the heightened immune activation seen in severe COVID-19 disease[9],[10]. Lastly, many chemotherapy and targeted therapies require high dose steroid premedication or therapy and need hospital visits for infusion, both of which predispose to infections.

The COVID-19 pandemic has caused a conundrum of problems specific to cancer patients such as increasing need for intensive care unit (ICU) admissions and ventilatory support; redeployment of resources resulting in delayed cancer care; suspension of clinical trials limiting availability of lifesaving therapies; delay in diagnostic and screening programs; modification of standardized protocols that might compromise cancer control; and reduced willingness among cancer patients to visit hospitals owing to the fear of infection[11,12]. The majority of published reports on cancer patients with COVID-19 have been single institutional retrospective studies with selective reporting of outcomes. There remain a multitude of unanswered questions regarding the actual of impact of COVID-19 on cancer patients such as differences in survival outcomes in patients with active cancer and cancer survivors; the impact of various oncologic therapies; difference in outcomes in subtypes of cancer; along with the safety and interaction of COVID-19-directed therapy with cancer-directed therapy. We performed this systematic review and meta-analysis to interrogate and summarize the lessons learned from the clinical reports on various malignancies that have reported mortality outcomes in cancer patients affected by COVID-19.

## Methods

This systematic review was performed according to the Preferred Reporting Items for Systematic Reviews and Meta-Analyses (PRISMA) recommendations[13]. The complete search protocol is provided in **Supplement 4**. Institutional review board approval was not required for this study since no patient (CRD42020186671).

## Data sources

A systematic electronic search was performed in PubMed/MEDLINE, Embase, Cochrane Central, Google Scholar, and MedRxiv databases to identify studies reporting outcomes on cancer patients with COVID-19 from December 1, 2019 to May 23, 2020. The Medical Subject Heading terms used for the search have been provided in **Supplement 5**. An independent review of the abstracts and full paper articles was performed by 2 reviewers (BV and VC). The duplicates were removed and the titles of articles were then evaluated. Abstracts found to be relevant to the topic of interest were shortlisted. Full-length papers of the shortlisted articles were assessed for the eligibility criteria. The articles that fulfilled the inclusion criteria were shortlisted for final systematic review. The included study references were cross-searched for additional studies. ClinicalTrials.gov, World health organization (WHO) International Clinical Trials Registry Platform (ICTRP) and Cochrane COVID registry were assessed for completed and ongoing

Reviews and Meta-Analyses (PRISMA) recommendations[13]. The complete search protocol is provided identifiers were disclosed. The systematic review has been registered in the PROSPERO database clinical trials related to cancer patients with COVID-19. The articles were reviewed independently by two authors (BV and VC) and any disagreement was resolved by consensus with a third author (SK). Reasons for excluding studies were documented.

## Study selection

The inclusion criteria were as follows: 1) studies reporting mortality outcomes in cancer patients with SARS-CoV-2 infection; 2) all types of studies (including randomized controlled trials (RCTs), prospective, retrospective and case series) comprising more than 10 patients; and 3) patients≥ age.

Exclusion criteria were: 1) Studies that did not report mortality outcomes in cancer patients with SARSCoV-2 infection; and 2) pre-clinical studies, epidemiological studies, autopsy series, incidence and prevalence studies, or news reports.

## Data extraction and quality assessment

The data was extracted by two authors independently into pre-defined forms. The following data was extracted from the studies: First author, mean age, study design, number of patients, gender, comorbidities, COVID-19-directed treatment, cancer subtypes, different treatments received, number of cancer patients and cancer survivors with corresponding mortality outcomes, ICU admissions, DNR/DNI numbers, need for mechanical ventilation, and progression to severe disease. Data for both cancer and non-cancer patients (for available studies) were extracted separately.

## Data synthesis

Percentages for categorical variables and medians with interquartile range (IQR) for continuous variables were presented. Pooled rates with 95% confidence intervals (CI) were calculated for individual arms. Odds ratios (OR) comparing cancer patients with non-cancer control patients was reported with 95% CI and *p* <value 0.05 was considered statistically significant. The random-effects model described by DerSimonian and Laird was used for analysis. Corresponding forest plots were constructed for both primary and secondary outcomes. Study heterogeneity was assessed using the inconsistency index(*I*^2^ statistic) with values of 0–30%, 31%–60%, 61%–75% and 76%–100% indicating low, moderate, substantial and considerable heterogeneity, respectively. All analyses were performed using statistical software Open Meta analyst (CEBM, Brown University, Rhode Island, USA) and Review Manager Version 5.3 (The Nordic Cochrane Centre, Copenhagen, Denmark). Sub-group analyses were performed for the following, when data was available: 1) subtypes of cancer; 2) type of cancer-directed therapy; 3) patients with active cancer versus cancer survivors; and 4) mortality outcome based on geographic location.

## Results

### Study search and study characteristics

The literature search resulted in 4,854 articles, of which 92 articles underwent full review and 31 were included in the final analysis (**Figure 1**). Among the studies included for systematic review, 26 were retrospective studies (**Supplementary data 1**) and 5 are ongoing clinical trials on cancer patients with no reported outcomes at the time of conducting this meta-analysis (**Supplementary data 2**). The data from 26 studies were included in the meta-analysis; 17 studies had multiple cancer types and 9 studies pertained to a single cancer type. Ten studies were performed in China, 6 in the United States, 3 in the United Kingdom, 3 in Italy, and 2 each in Spain and France.

**Figure 1:**
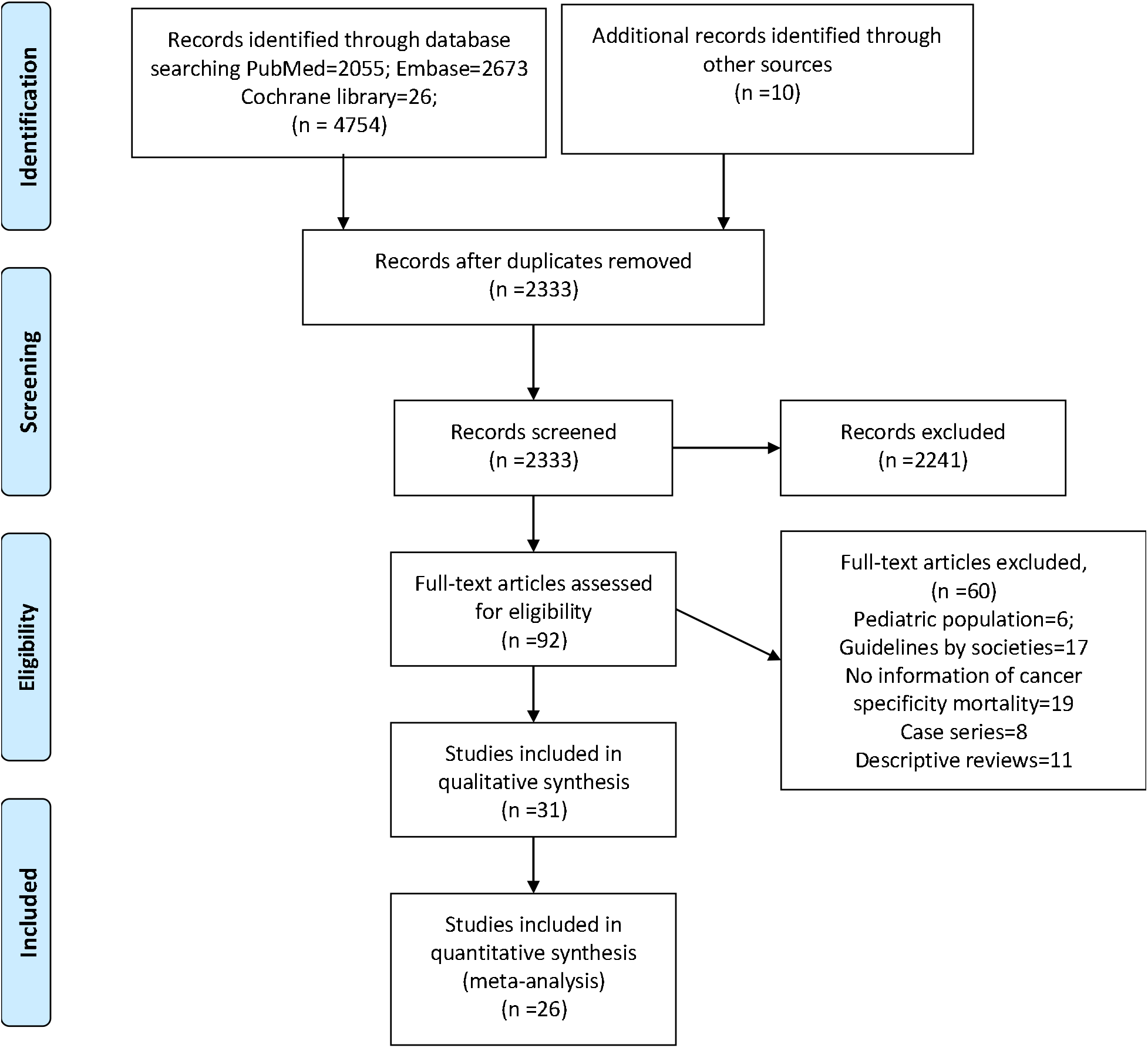
PRISMA flow diagram of the selection of studies to be included in the systematic review and meta-analysis.

### Patient characteristics

There were 23,736 cancer patients (mean age 65.1 ± 8.02 years) versus 157,587 non-cancer patients (50.3 ± 11.9 years). The proportion of males were 52.6% (n = 1,413) versus 52.9% (n = 9,067), respectively. The proportion of comorbidities was higher in the cancer arm with the prevalence of smoking being 31.1%; hypertension 46.6%; diabetes mellitus 20.4%; cardiac disease 34.9%; cerebrovascular disease 9.1%; chronic liver disease 9.2 %; chronic kidney disease 10.8%; and chronic lung disease 14.7% compared to the non-cancer arm with prevalence of smoking being 7.3%; hypertension 24.2%; diabetes mellitus 5.4%; software Open Meta analyst (CEBM, Brown University, Rhode Island, USA) and Review Manager for the following, when data was available: 1) subtypes of cancer; 2) type of cancer-directed therapy; 3) patients with active cancer versus cancer survivors; and 4) mortality outcome based on geographic cardiac disease 7.3%; cerebrovascular disease 3.9%; chronic liver disease 6.5 %; and chronic kidney disease 4.1%.The demographics are provided in **Supplementary data 3**.

### Primary outcomes reported in the studies

Twenty-six studies provided data on cancer patient mortality. The pooled all-cause in-hospital mortality rate was 19.2% (95% CI: 14.9% – 23.5%) (n = 23,736) with a median follow up duration of 45 days (range of 21–104 days). Comparing the mortality between the cancer and non-cancer patients, 10 studies (n = 165,980) provided such data, with a pooled rate of 16.6% (95% CI: 10.4% – 22.8%) and 5.4% (95% CI:4.1% – 6.7%), respectively (odds ratio (OR) 2.54, 95% CI: 1.47 – 4.42, I^2^ = 92%, p = 0.00009). Further sub-group analysis performed on studies with sample size over 100 confirmed the aforementioned findings (OR 2.17, 95% CI: 1.16 – 4.07, I^2^ = 95%, p = 0.02).

In subgroup analysis stratifying based on geographical location, cancer patients in China had higher mortality than non-cancer patients (OR 6.62, 95% CI: 2.68 – 16.34, I^2^ = 59%, p = 0.001), but this trend was not observed in the United States (OR 1.56, 95% CI: 0.62 –3.96, I^2^ = 95%, p = 0.35) or Europe (OR 1.69, 95% CI: 0.81 – 3.52, I^2^ = 89%, p = 0.161).

Seventeen studies reported ICU admission rates in cancer patients, with a pooled ICU admission rate of 12.6% (95% CI: 8.9%-16.3%) (n = 1,834). When evaluating the 3 studies (n = 11,587) which also provided information on the non-cancer population, the pooled ICU admission rates in the respective cohorts were 12.6% (95% CI: 4.7% – 20.5%) and 7.1% (95% CI: 5.1% – 9.1%), with no significant difference between the two groups (OR 2.18, 95% CI: 0.78 – 6.04, I^2^ = 85%, p = 0.13).

Fifteen studies reported the need for mechanical ventilation in cancer patients, with a pooled intubation rate of 10.9% (95% CI: 6.8% – 15.0%) (n = 1,813). When examining the 3 studies (n = 6,353) which also provided information on non-cancer cases, the pooled rates were 10.8% (95% CI: 7.9% – 13.7%) and 4.9% (95% CI: 2% – 7.8%), respectively (OR 2.43, 95% CI: 1.43–3.88, I^2^ = 19%, p = 0.0002).

### Cancer subtype specific outcomes

The most common type of cancer reported among COVID-19 patients were hematological malignancies with a reported pooled proportion of 34.3% (95% CI: 7.4% – 61.3%) (n = 2,316). This was followed by breast cancers at 29.2% (95% CI: 6.1% – 51.40%) (n = 1,945), lung cancers 23.7% (95% CI: 2.0% –45.3%) (n = 2,051), gastrointestinal malignancies 15.2% (95% CI: 11.7% – 18.7%) (n = 2643), prostate cancers 11.1% (95% CI: 5.7% – 16.6%) (n = 2039), gynecological cancers 9.6% (95% CI: 5.7% – 13.5%) (n = 1077), head and neck cancers 3.7% (95% CI: 2.4% – 5.0%) (n = 883), brain tumors 3% (95% CI: 0.8%-5.3%) (n = 465), and other cancers 2.63% (95% CI: 0.7% – 5.19%) (n = 2033). Hematological malignancies had the highest pooled all-cause in-hospital mortality rate of 33.1% (95% CI: 16.1–50.1%) (n = 266) followed by lung cancer at 28% (95% CI: 18.8–37.1%) (n = 161), gastrointestinal malignancies at 19.8% (95% CI: 6.3–33.3%) (n = 99), and breast cancer at 10.9% (95% CI: 3.5%-18.3%) (n= 193). The numbers reported in the other malignancies were insufficient to perform a subset analysis.

We performed an additional subgroup analysis for mortality by stratifying hematological malignancies vs. other malignancies, lung vs. other malignancies, gastrointestinal malignancies vs. other and breast vs. other malignancies. Hematological malignancies had the highest OR of death (2.39, 95% CI: 1.17 – 4.87, I^2^ = 49%, p = 0.02) (n = 878) followed by lung cancer (1.83, 95% CI: 1.00 – 3.37, I^2^ = 19%, p = 0.05) (n = 646), which were both statistically significant; the remainder were not (p>0.05 for both).

### Treatment related outcomes

The most common treatment modality reported in cancer patients affected with COVID-19 was chemotherapy (pooled rate of 30.3%, 95% CI: 22.3%-37.8%) (n = 1,166) followed by hormonal therapy(17.4%, 95% CI: 6.9%-27.9%) (n = 332), targeted therapy 15.4% (95% CI: 9.5%-21.2%) (n = 837), radiotherapy 13.8% (95% CI: 7%-20.7%) (n = 790), immunotherapy 9.1% (95% CI: 5.2%-12.9%) (n = 1345) and surgery 7.3% (95% CI: 5.2%-9.4%) (n = 776). Subgroup analysis of treatment modalities showed no significant differences in mortality associated with radiotherapy (OR 0.72, 95% CI: 0.36 –1.42, I^2^ = 0%, p = 0.34) (n = 335), chemotherapy (OR 0.74, 95% CI: 0.40 – 1.39, I^2^= 0%, p = 0.35) (n = 386), immunotherapy (OR 1.61, 95% CI: 0.65 – 4.00, I^2^ = 30%, p = 0.31) (n = 467) and targeted therapy (OR 2.57, 95% CI: 0.93 – 7.09, I^2^ = 0%, p = 0.07) (n = 181). The number of reported patients having undergone hormonal therapy and surgery were not adequate for subgroup analysis.

### Cancer survivors

The definition of cancer survivors was defined variably by different studies. Wang et al. defined cancer survivors as patients treated for cancer within the past 5 years without any active disease [14]. Zhang et al. defined cancer survivors as patients who had not received cancer-directed therapy in the in the past one month[15]. Liang et al. defined cancer survivors as patients who had not received cancer-directed therapy recently[16]. Patients receiving active cancer-directed therapy were at higher risk of developing severe disease compared to patients classified as cancer survivors with OR 2.58 (95% CI: 1.35 – 4.95, I^2^ = 0%, p = 0.004) (n = 366).

### Ongoing cancer-directed therapy clinical trials in COVID-19 patients

There are currently only 5 active ongoing clinical trials that are assessing interventions specific to cancer patients. This represents only 0.7% of all the ongoing treatment based studies[17]. The interventions being assessed include the chloroquine analog (GNS561), an anti PD-1 antibody (nivolumab); an anti-interleukin-6 receptor (tocilizumab); an anti-interleukin-8 (BMS-986253), a SUMOylation inhibitor (TAK-981), and azithromycin. The chloroquine analog and azithromycin are being tested as prophylactic agents whereas the others are being evaluated as therapeutic agents (supplementary data 2).

## Discussion

This systematic review and meta-analysis of 181,323 patients from 26 studies involving 23,736 cancer patients is the largest meta-analysis to the best of our knowledge assessing outcomes in cancer patients affected by COVID-19. Our meta-analysis shows that cancer patients with COVID-19 have an increased likelihood of death compared to non-cancer COVID-19 patients, which was particularly driven by patients in China. Cancer patients were more likely to be intubated than non-cancer patients but ICU admission rates were not statistically significant between the two groups. Hematological malignancies were associated with the highest mortality, followed by lung cancer. There was no association between receipt of a particular type of oncologic therapy and mortality.

Some of our findings are rather intuitive; nonetheless, having objective data to confirm our suspicions is helpful in making evidence-based recommendations for deployment of limited resources within healthcare environments. For instance, the higher mortality in patients with hematological malignancies could be readily explained by the greater degree of immunosuppression utilized in the treatment of these patients, even in the absence of bone marrow or stem cell transplantation. The underlying immunosuppressive microenvironment due to dysfunctional immune cell production is a fundamental attribute of the malignancies that drives their pathogenesis[18]. Faced with an added insult (e.g. COVID-19), these patients may not have adequate immune reserves to combat the infection. In a flow cytometric analysis of 522 patients from China, T-cell lymphopenia was an important prognostic factor for mortality in COVID-19 patients[19]. The OpenSAFELY Collaborative from United Kingdom published one of the largest epidemiological reports, stating that the hazard ratio (HR) of death in patients with hematological malignancies was 3.52 (2.41–5.14) as compared to 1.56 (1.29–1.89) for solid tumors[7].

Our investigation did not show any clear association between treatment modality and mortality, which is reassuring. There have been concerns regarding an association between immune checkpoint inhibitors and the hyperactive phase of COVID-19 infection. In an analysis of 67 consecutive lung cancer patients with COVID-19 treated with PD-1 blockade, no significant association was found between the timing of PD-1 blockade and COVID-19 severity as well as mortality[9]. We observed a numerical increase in the odds of death from targeted therapy and chemotherapy, but it was not statistically significant. However, there was a lack of information regarding the particular agent/class; hence, we were not able to perform corresponding subgroup analyses. Of note, no association was found between radiation delivery and COVID-19. Radiation is known to deplete the circulating as well as resident lymphocyte subpopulations and this lymphopenia is known to last months to years[8]. However, these results should also be interpreted with caution in view of the lack of robust numbers in the groups analyzed.

A few caveats about our analysis are noteworthy. The pooled mortality rates in such a meta-analysis may be misleading given that cancer patients are often older and have more comorbidities. Hence, the actual magnitude of mortality in cancer patients with COVID-19 using age-matched cohorts might be lower than reported in these studies. Additionally, mortality differences seemed to be driven by Chinese patients, which could imply unforeseen COVID-19-treatment-related effects or genetic polymorphisms as compared to Western populations. Within Western populations, the findings of our analysis may still help inform how resources are redeployed within oncology units or cancer centers. For instance, the greater mortality among patients with hematological malignancies may argue for marshalling additional resources to these units in a cancer center. It may also support a conscious decision to delay highly immunosuppressive treatment such as bone marrow transplantation, stem cell transplantation or CAR-T cell therapy. However, modifying standard therapeutic protocols to accommodate predicted disparities in mortality engenders additional risks in terms of disease progression. Clearly, such modifications should be guided by a nuanced risk-benefit analysis based on the best available data.

## Limitations of the study

As is customary with such data, the results should be interpreted cautiously, as the studies were heterogeneous and sample sizes were variable. The majority of studies were single-institutional series with selective reporting of data and high publication bias. The contribution of age, co-morbidities, preexisting immunodeficiency, and polypharmacy to mortality outcomes in cancer patients with COVID-19 remain difficult to assess. Furthermore, none of the studies reported mortality outcomes in relation to COVID-19-directed therapy. Accordingly, the potential interactions between cancer-directed therapy and COVID-19-directed therapy were not documented in any of the studies[20]. Taken together, these attributes make the conclusions arising from our meta-analysis predominately hypothesis-generating rather than definitive and/or conclusive. However, at this point in time, these studies remain the core of the limited evidence on this topic to date, and higher quality of studies in the future may allow for corresponding improvements to future meta-analyses.

## Implications for clinical practice

Cancer patients with COVID-19 have a higher probability of severe disease, increased ventilatory requirements, and mortality compared to the general population. Patients with hematological malignancies and lung cancer are at increased risk of death compared to other subtypes of cancer. There is a need for prospective registration of cancer patients with COVID-19 in registry initiatives like the UK Coronavirus Cancer Monitoring Project (UKCCMP) and ASCO Survey on COVID-19 in Oncology Registry (ASCO Registry) for better understanding of the impact of cancer-directed therapies as well as COVID-directed therapies on mortality and morbidity outcomes[21].

## Data Availability

Data will be avaliable on request to the corresponding author

## Acknowledgments

We thank the researchers who posted the studies in the public domain.

## Figure legends

**Figure 2:**
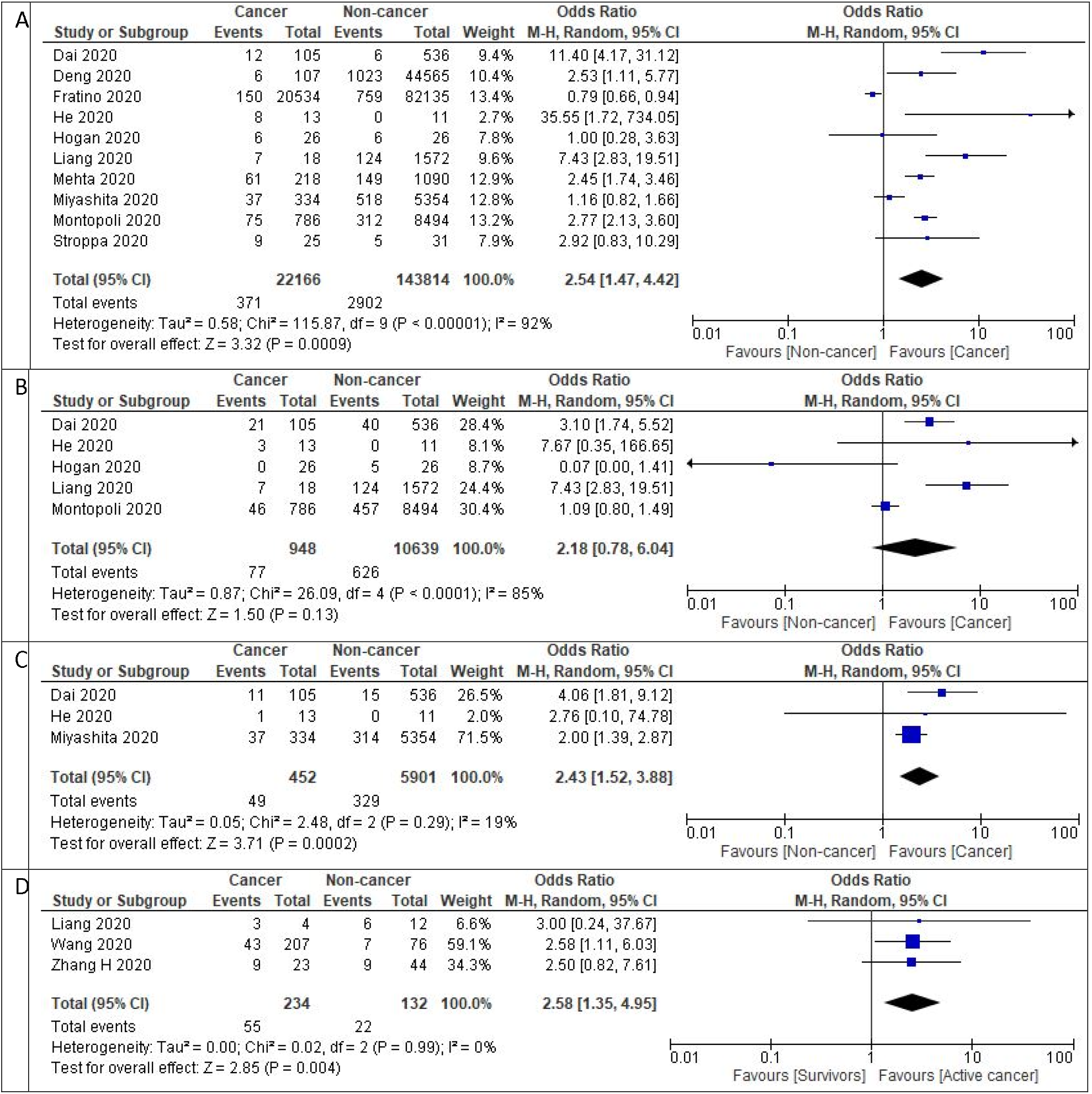
Prognosis of patients with COVID-19 1 A) Forest plot of pooled in-hospital all-cause mortality rates between cancer patients and non-cancer patients.1B) Forest plot of ICU admission rates between cancer patients and non-cancer patients.1C) Forest plot of intubation rates between cancer patients and non-cancer patients.1D) Forest plot of severe disease between active cancer patients and cancer survivors. Odds ratio calculated using the Mantel-Haenszel random-effects model.

**Figure 3:**
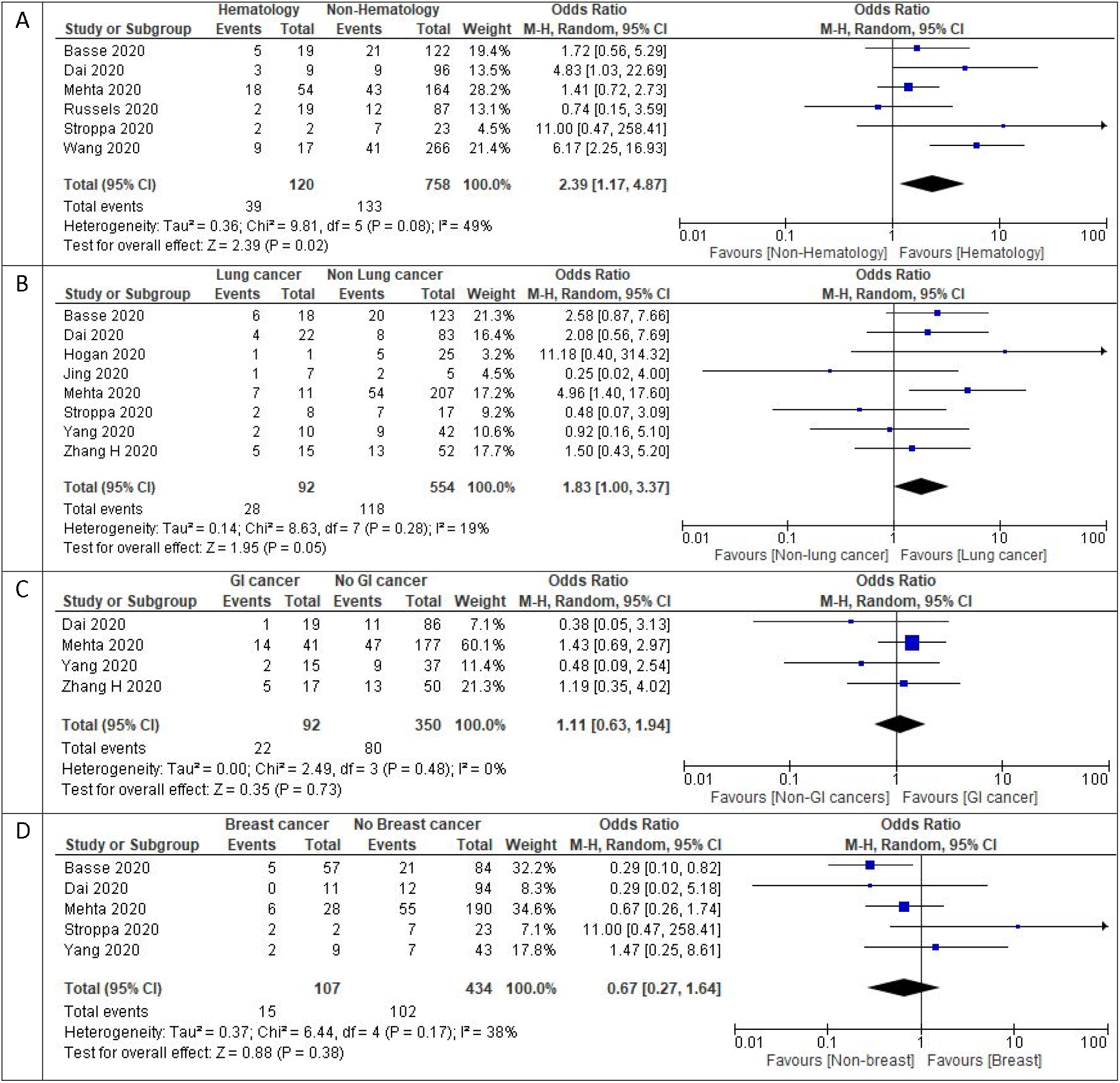
Mortality outcomes in cancer subtypes with COVID-19 1 A) Forest plot of pooled in-hospital all-cause mortality rates between hematological cancer patients and non-hematological cancer patients.1B) Forest plot of pooled in-hospital all-cause mortality rates between lung cancer patients and non-lung cancer patients 1C) Forest plot of pooled in-hospital all-cause mortality rates between gastrointestinal cancer patients and non-gastrointestinal cancer patients.1D) Forest plot of in-hospital all-cause mortality rates between breast cancer patients and non-breast cancer patients. Odds ratio calculated using the Mantel-Haenszel random-effects model.

**Figure 4:**
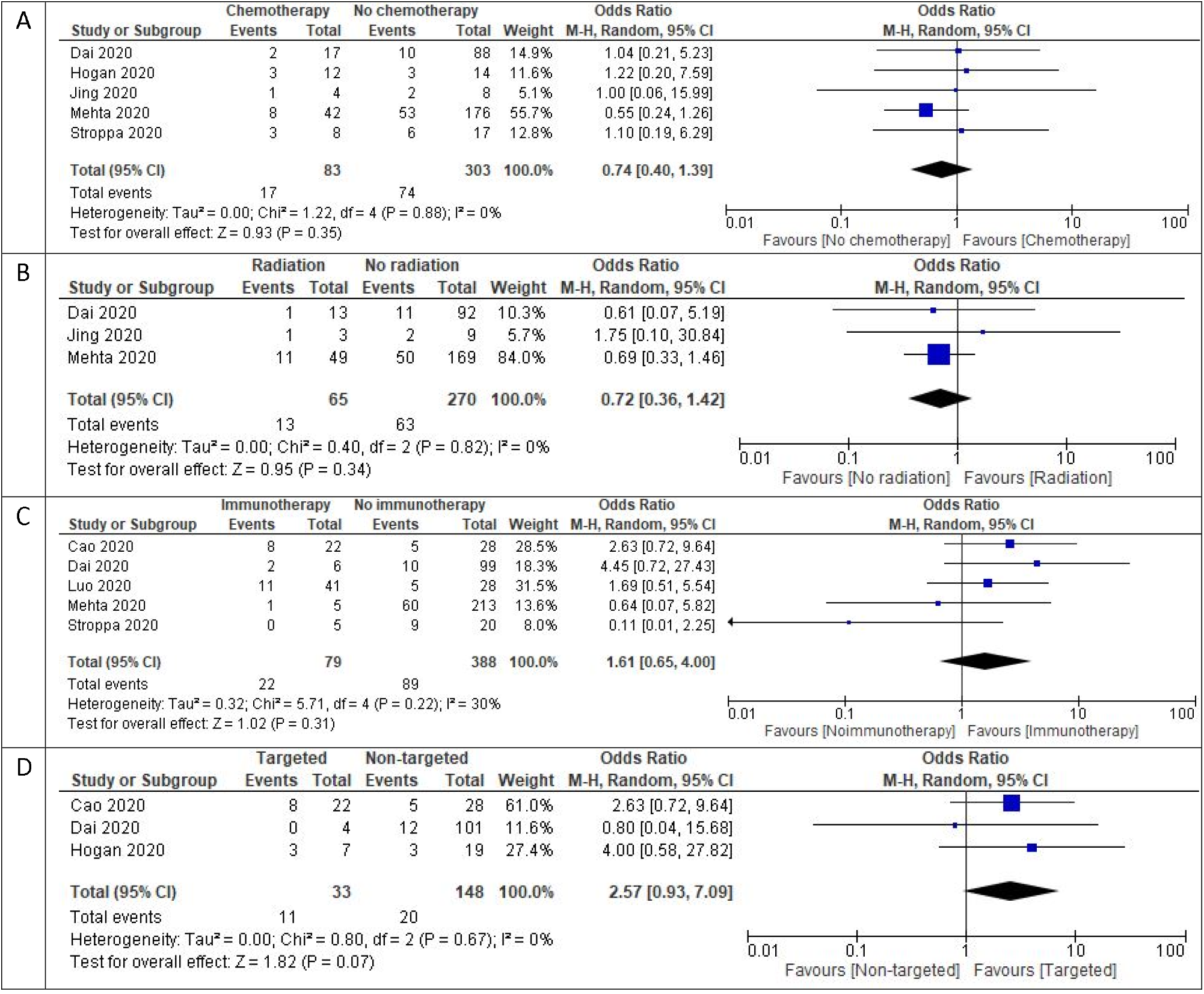
Mortality outcomes with different types of treatment with COVID-19 1 A) Forest plot of pooled in-hospital all-cause mortality rates between patients receiving chemotherapy vs. Other modalities 1B) Forest plot of pooled in-hospital all-cause mortality rates between patients receiving radiotherapy vs. other modalities 1C) Forest plot of pooled in-hospital all-cause mortality rates between patients receiving immunotherapy vs. other modalities 1D) Forest plot of pooled-in hospital all-cause mortality rates between patients receiving targeted therapies vs. other modalities. Odds ratio calculated using the Mantel-Haenszel random-effects model.

